# Neural signatures and personalized neuromodulation in a subject experiencing context-dependent inhibitory control deficits

**DOI:** 10.1101/2025.06.06.25328536

**Authors:** Layth S. Mattar, Shraddha Shah, Lily S. Chamakura, Denise Oswalt, Yue Zhang, Davin Devara, Jung Uk Kang, Zahra Jourahmad, Ryan Jafri, Geoffrey Liu, Joshua Adkinson, Isabel A. Danstrom, Xiaoxu Fan, Yvonne Y. Reed, Kelly R. Bijanki, Alica Goldman, Lu Lin, Vaishnav Krishnan, Nicole R. Provenza, Andrew J. Watrous, Sarah R. Heilbronner, Sameer A. Sheth, Garrett P. Banks, Eleonora Bartoli

## Abstract

The ability to override prepotent actions is critical to control impulses and adjust behavior depending on goals and contextual needs. In this study, we investigate the inhibitory control abilities of a patient diagnosed with Klüver-Bucy Syndrome following a left temporal resection. The patient presented with disruptive hypersexuality symptoms akin to compulsions, leading to the inability to control and suppress inappropriate actions. The patient was recruited for the current research study while undergoing intracranial monitoring for epilepsy, to investigate the cognitive and neural processes underlying the patient’s inhibitory control symptoms. We formulated the hypothesis that an inhibitory control deficit emerges in response to provocative triggers, and we designed a personalized paradigm pairing arousing images with a classic inhibitory control task. We not only confirmed disrupted performance following exposure to triggering, provocative material, but we also leveraged the simultaneously recorded neural data to identify a biomarker reflecting inhibitory control failures. Next, we repeated the experimental paradigm during and after personalized neuromodulation via direct high-frequency stimulation of the right inferior frontal cortex. The patient displayed a marked improvement in his behavior during neuromodulation, mirrored by changes in neural activity, spanning spectral features, event-related potentials and functional connectivity. Altogether, our study revealed that the patient’s symptoms were not due to a global inhibition deficit, but to a specific control issue triggered by exposure to provocative material. Overall, our work showcases a feasible, effective approach towards data-driven personalized neuromodulation, which could be leveraged to mitigate specific inhibitory control deficits and potentially other symptoms of executive dysfunction.

*Graphical abstract. This case study investigates context-dependent inhibitory control abilities in a patient with Kluver-Bucy syndrome during intracranial monitoring for epilepsy*.

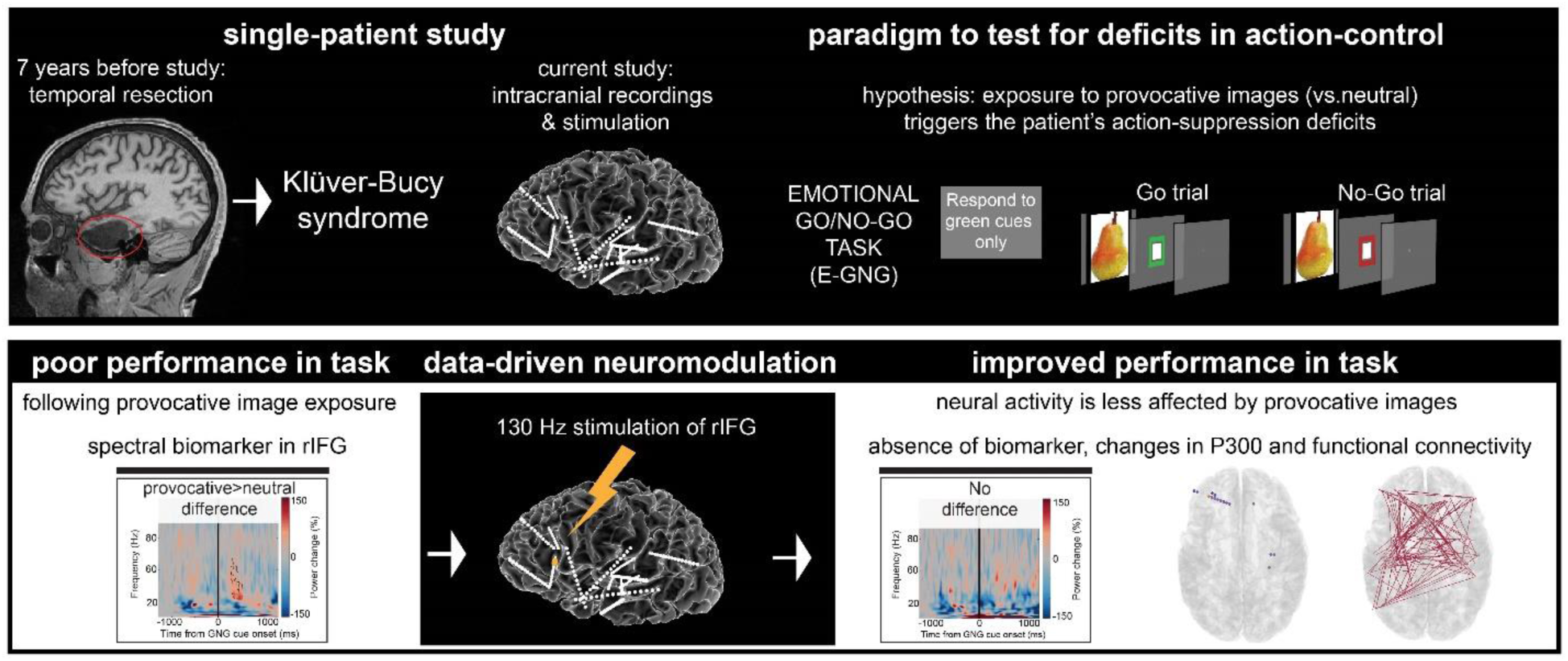

## 1. Introduction

This case study analyzes the neural and behavioral activity, and the influence of neuromodulation upon said activity, of an individual with a diagnosis of epilepsy and Klüver-Bucy Syndrome (KBS). KBS, a neuropsychiatric disorder associated with temporal lobe lesions, is diagnosed by having at least three of the following seven symptoms: hyperorality, hypermetamorphosis, hypersexuality, bulimia, placidity, visual agnosia, and amnesia (Clay et al., 2019). The patient developed symptoms compatible with KBS following a left temporal resection for drug-resistant epilepsy, seven years prior to the current study. Postoperatively the patient continued to suffer from seizures and developed an atypical presentation of KBS with hyperorality and hypersexuality as his primary symptoms. The patient likened the occurrences of hypersexuality akin to compulsions, which led to significant distress in his life. These compulsions presented as an inability to refrain from actions leading to inappropriate sexual behaviors, especially when exposed to triggering situations or materials.

Cognitive and emotional changes have been observed following temporal lobe damage, whether resulting from disease-related lesions or surgical treatment to manage conditions such as epilepsy. Most commonly, these changes manifest as verbal and general memory decline, but can also appear as more niche but equally disruptive symptoms such as hypersexuality (Baird et al., 2002; Hamberger and Drake, 2006). The precise source of these changes is unclear, and may involve aberrant activity within the limbic system (as in classic KBS) and/or pathology to the temporal lobe contralateral to the site of damage (Baird et al., 2007).

The patient was recruited for the current study while undergoing invasive monitoring of epileptic activity through stereo-electroencephalography (sEEG). The atypical presentation of their symptoms with respect to classic KBS, their surgical history, and current literature studying similar cases and phenomena (Caro and Jimenez, 2016; Lanska, 2018) led us to formulate the hypothesis of a context-specific inhibitory control deficit, namely the inability to refrain from inappropriate behaviors following exposure to sexually provoking stimuli. Attempts to determine the neural circuit behind the restraint of prepotent behaviors have led to a variety of models (Gavazzi et al., 2023; Wessel, 2023; Zhang et al., 2017), with most models including the right inferior frontal gyrus (rIFG) as a key component responsible for initiating action stopping (Aron, 2011; Bartoli et al., 2018; Cai et al., 2017a; Choo et al., 2022; Kang et al., 2025), signaling the need to interrupt ongoing motor plans to the basal ganglia-motor cortex pathway via the subthalamic nucleus.

The influence of contextual factors over inhibitory control abilities can be assessed by combining action suppression paradigms (e.g., go/no-go and stop-signal tasks) with emotional images (Albert et al., 2010; Battaglia et al., 2021; Mirabella et al., 2024; Zhao et al., 2019). Both positive and negative emotions can interfere with action-stopping functions, although the effects are quite limited in healthy individuals and depend on the type of inhibition (Littman and Takács, 2017a; Montalti and Mirabella, 2023). The emotional interference effect in the stop-signal task is reflected by an increased stop-signal reaction time (SSRT), and it has been interpreted as the attentional capture effect of high-arousing stimuli, regardless of whether the stimulus is negative or positive (Verbruggen and De Houwer, 2007). Provocative images, such as erotic pictures, have also been shown to slow the SSRT (Yu et al., 2012), but the effect is abolished if participants think they are being monitored (Yu et al., 2015), demonstrating volitional control over the degree of attentional capture. Go/no-go paradigms, measuring reactive control, are more robust to emotional interference, with no effects on accuracy (Albert et al., 2010; Littman and Takács, 2017a; Zhao et al., 2019), although neural measures, like event-related potentials (ERP), can reveal sensitivity to the emotional interaction in the absence of behavioral effects (Albert et al., 2010; Zhao et al., 2019). This evidence indicates that emotional content can capture resources and slow down action-stopping, especially in the stop-signal task, while action withholding in go/no-go does not typically suffer from contextual effects.

In this case study, we hypothesized that the general ability to suppress a motor response would be intact, but that the exposure to provocative images would cause a profound interference and impair the patient’s inhibitory control abilities in a context-dependent manner. We collected neural data from the most responsive regions during the presentation of emotional visual stimuli to analyze the relationship between pathological neural activity and the subject’s behavior, as has been seen in other psychiatric and neurological conditions (Arnold et al., 2012; Erk et al., 2010; García-García et al., 2016; Soleimani et al., 2025). Additionally, we aimed to improve the patient’s control abilities with personalized neuromodulation. Studies have repeatedly found that targeted deep brain stimulation (DBS), if accurately placed and appropriately configured, can lead to significant neuropsychological alterations in other nodes of the same circuit, and influence the behavioral outcomes of that pathway (Bartoli et al., 2024; Harmsen et al., 2022; Kundu et al., 2018; Neumann et al., 2023). These findings suggest that electrical neuromodulation in critical inhibition nodes like the rIFG, or other functionally relevant regions, may mitigate the inhibition deficiency and improve action-stopping (Wessel et al., 2013a). Furthermore, recent efforts on addiction and substance abuse disorders demonstrated the efficacy of combining neuromodulation methods with patient-specific exposure/cognitive paradigms, enhancing therapeutic outcomes (Sathappan et al., 2019; Soleimani et al., 2025; Spagnolo et al., 2020).

To this end, we performed a series of experiments to assess the interaction between inhibitory control and arousing, provocative stimuli designed to trigger the subject’s hypersexuality. Experiments were performed with and without targeted neuromodulation. Specifically, intracranial neural and behavioral data were collected during an emotional go/no-go task (E-GNG), with provocative and neutral visual stimuli presented before the go and no-go cues. We reasoned that, in an E-GNG task, only a profound, pathological interference would be capable of a behaviorally-measurable effect, as this paradigm is generally robust to emotional interference effects (Albert et al., 2010; Littman and Takács, 2017a; Mancini et al., 2022; Zhao et al., 2019). Next, we combined a data-driven approach with our hypothesis regarding inhibitory control dysfunction, to find a targetable biomarker for neurostimulation: a location displaying an increased response to the provocative images. Our analysis revealed rIFG to be highly responsive, which we then targeted with direct, high-frequency stimulation to determine if patient performance could be improved with personalized neuromodulation. For all experiments, we analyzed the patient’s behavioral performance and neural activity, focusing on spectral changes and on a well-established event-related potential in GNG paradigms, the P300 (Bruin and Wijers, 2002), measured intracranially. We further analyzed functional connectivity using mutual information to evaluate the effect of neuromodulation on large-scale interareal dynamics.

## 2. Methods

### 2.1 Human Subject

One male patients in his sixties with a history of drug-resistant epilepsy provided informed consent to participate in this study during intracranial epilepsy monitoring via sEEG at Baylor St. Luke’s Medical Center (Houston, Texas, USA). The subject had previously undergone a left temporal resection seven years prior to the current study for medically refractory epilepsy (Figure 1A) at a different institution. The patient did not attain seizure freedom following the resection and developed behavioral changes leading to a diagnosis of KBS which included hypersexuality, hyperorality, coprophagia, and urophagia. The intracranial monitoring at our site was performed as part of the clinical care for seizure onset zone determination, seven years after the temporal resection, and the subject was recruited to participate in the research activities during his stay. Experimental procedures were approved by the Institutional Review Board at Baylor College of Medicine (IRB protocol number H-18112).

**Figure 1.**
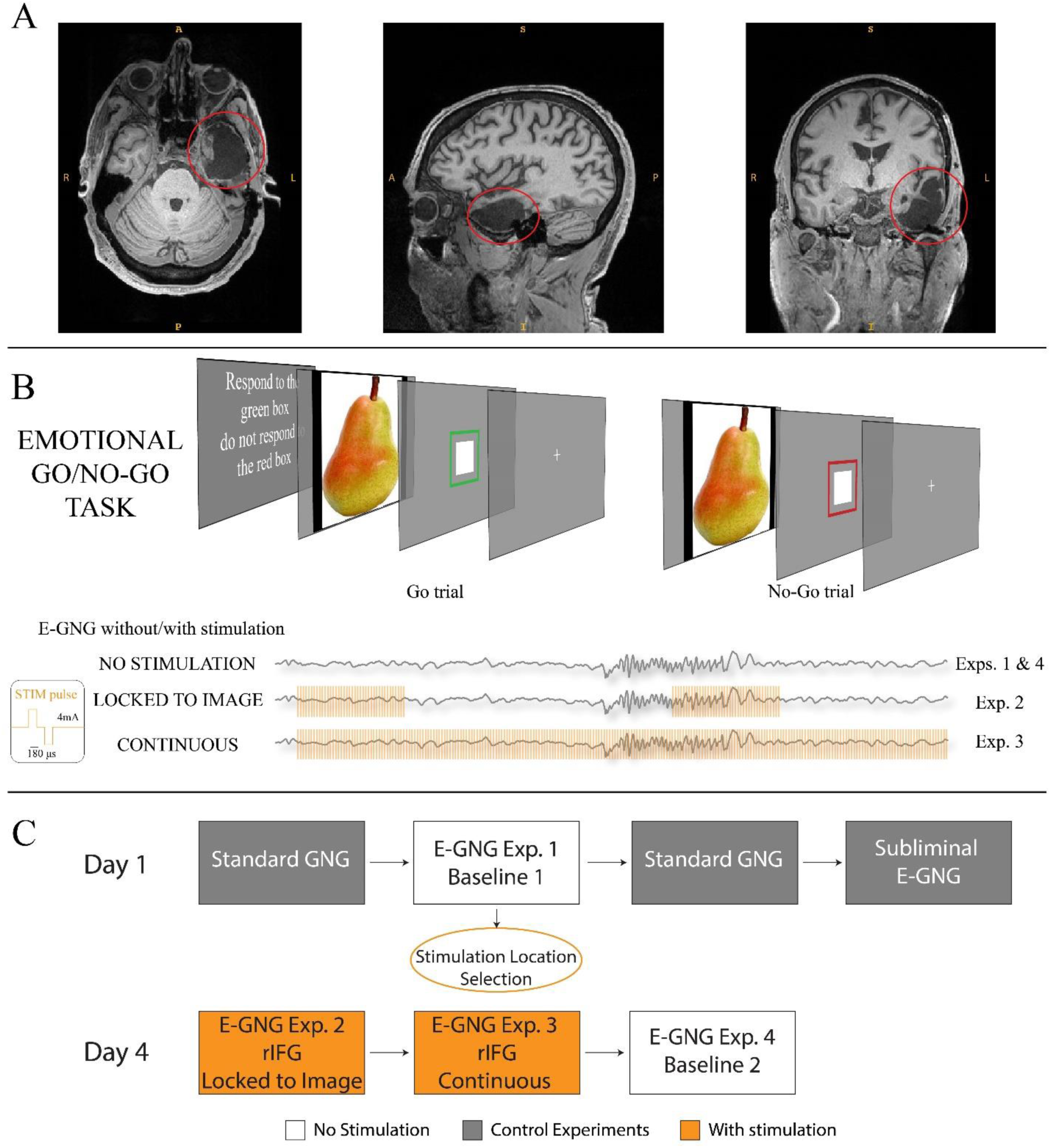
Task and experimental design. A) MRI axial, sagittal, and coronal views, showcasing the left temporal resection performed seven years prior to the current study (highlighted in red). B) The E-GNG task consisted of alternating blocks that included neutral or provocative task-irrelevant content displayed prior to the GNG cues. A depiction of the different E-GNG stimulation parameters is shown at the bottom of the panel: baseline experiments are performed without stimulation (Exp.1 and 4), and stimulation experiments entailed stimulation locked to the image presentation (Exp.2) or continuous stimulation (Exp.3). C) Schematic of the experimental timeline. Control experiments and Exp.1 (baseline 1) were run first to determine the presence of an underlying general inhibition deficit and guide stimulation location parameters. Exp.2-4 were run three days later after analysis of Day 1 experiments.

### 2.2 Intracranial Electrodes

The subject underwent invasive surgery for the implantation of 16 depth probes with a combined total of 232 electrode contacts. The probes were placed bilaterally and in multiple anatomical locations across frontal and temporal regions. Depth probes had between 8 and 16 contacts with a 0.8 mm diameter and 3.5 mm center-to-center distance (PMT Corporation, MN, USA).

### 2.3 Electrode Localization

Electrode contact location was determined through the intracranial Electrode Visualization software pipeline, iELVis (Groppe et al., 2017). The post-op clinical CT images were co-registered with a pre-op T1 anatomical MRI via the linear image registration tool (FLIRT) from the Functional Magnetic Resonance Imaging for the Brain Software Library (FMRIB) (Jenkinson and Smith, 2001). Electrode contacts were then manually identified on the generated CT-MRI overlay in BioImage Suite (Papademetris et al., 2006). The *xyz* coordinates of each electrode translate to their position in space (mm) in an R-A-S (right-anterior-superior) convention with an origin at the central voxel of the T1 volume. These coordinates were used for anatomical labeling purposes. An expert neuroanatomist (S.R.H.) labeled the electrode contact locations, and for contacts deep in white matter, the XTRACT toolbox (Warrington et al., 2020) was used for additional guidance.

### 2.4 GNG Tasks

We employed two different GNG tasks: a standard version (GNG) and an emotional version (E-GNG). The standard version was run first as a control, followed by the first E-GNG experiment, and the standard was run again as a second control. A subliminal version of the E-GNG was run as a third control. The E-GNG was then paired with different stimulation protocols. We first describe the overall structure for both tasks and then describe the experimental procedure.

#### 2.4.1 Standard GNG

First, we employed a short standard GNG task to familiarize and train the subject with the paradigm. The participant was instructed to deliver a fast response to a go-cue (black square with green outline, subtending 4.3 degrees of visual angles) by pressing the left arrow on a handheld keyboard and to withhold the response when a no-go was presented (black square with red outline, 30% of trials). Each trial started with a fixation cross (500 ms) followed by the cue (2000 ms) and then by a 500 ms inter-trial interval (blank screen). The trials were organized in blocks of 50 trials (35 go, 15 no-go per block), with an initial training block consisting of 10 trials (not analyzed, used to familiarize the patient with the experimental procedure). After every block of 50 trials the patient was given a performance summary lasting 15 seconds outlining their accuracy, mean reaction time, and the option to take a small break.

#### 2.4.2 E-GNG

The E-GNG task followed the same basic structure of the GNG task described above, with the addition of task-irrelevant visual stimuli preceding the presentation of the GNG cues (Figure 1B). Each trial started with a fixation cross (500 ms) followed by a visual stimulus (1000 ms, or 50 ms in the control “subliminal” version), the cue (1000 ms), and then by a 500 ms inter-trial interval (blank screen). The visual stimuli fell into two groups, neutral and provocative images (500 x 500 pixels, subtending 15 degrees) presented against a dark gray background before the GNG cue. The provocative images were personalized based on the patient’s input and consisted of women in lingerie. The goal was to provide triggers similar to those that initiated hypersexual symptoms in the patient. The neutral images were made up of everyday objects (i.e., furniture and food). The E-GNG trials were organized in blocks of 50 trials, and each block contained exclusively neutral or provocative images (alternating blocks), with an initial training block consisting of 10 neutral trials (not analyzed). The GNG ratio was the same as for the standard task (30% no-go cues) and the subject was explicitly told to focus on the GNG cues and to try to ignore the images. We selected a blocked-design to exacerbate the effect of provocative images and their anticipation.

### 2.5 Procedure

#### 2.5.1 First E-GNG baseline and control experiments

The patient completed the experiments while seated on the hospital bed. The stimuli (fixation cross, visual images, and go/no-go cues) were displayed on a 50 inch TV approximately 246 cm away, and a response pad was used to record reaction times.

The standard GNG task was run first (200 trials, divided in four blocks). After that, the first E-GNG task was run (300 trials, divided in six blocks, alternating neutral and provocative blocks). Due to the presence of a strong behavioral effect, we asked the patient to perform the standard GNG task again (100 trials, two blocks) as a control. We then asked the participant to perform a “subliminal” version of the E-GNG task (visual images displayed for 50 ms, 300 trials, divided in six blocks, alternating neutral and provocative blocks) as a second control. Following these controls, which were used to assess behavior, we moved forward with the rest of the experimental design: we used the data recorded in the first E-GNG task to guide the neuromodulation experiments, performed three days later and described in the next section (Figure 1C). Only behavioral data, no neural data, was analyzed for control purposes from the standard GNG and subliminal E-GNG experiments.

#### 2.5.2 E-GNG experiments with/without neuromodulation

The subject completed the E-GNG paradigm a total of four times under two different neuromodulation parameters (Note: we consider the subliminal version a control, not counted here). Experiment 1: first baseline (no stimulation, 300 trials, described above). Neural recordings from Exp.1 were analyzed and used to select the stimulation location for the stimulation experiments (see below), which occurred three days later. All experiments had the same overall design but differed depending on the presence/settings of neuromodulation. Experiment 2: stimulation of the rIFG time-locked to image presentation (200 trials; stimulation on during each 1-second presentation of the images, both neutral and provocative). Experiment 3: continuous stimulation of the rIFG (200 trials; stimulation on for the whole duration of the experiment). Experiment 4: second baseline (no stimulation, 100 trials). Experiments 2-4 were performed on the same day and separated by a minimum of 30 minutes.

### 2.6 Stimulation parameters

For the stimulation experiments, bipolar stimulation was delivered using a CereStim R96 stimulator (Blackrock Microsystems, Utah, USA). Stimulation trains consisted of cathodic biphasic pulses (180 μs pulse width, 100 μs interphase gap) at a frequency of 130 Hz and with an amplitude of 4 mA. One region was stimulated: the rIFG (Exp.2-3). This region was not in the suspected seizure onset zone and all stimulation experiments were performed following an established pipeline to evaluate the safety of the stimulation targets and parameters with respect to epileptiform activity (Goldstein et al., 2019). In brief, the contacts selected for stimulation were evaluated by the attending epileptologist following an established procedure using trains of stimulation pulses at increasing amplitudes up to the target level (here, 4 mA at 130 Hz). This procedure reflects our standard pipeline to exclude any contacts that display an epileptiform after discharge, or contacts that generate subjective reports of any paresthesia, mood, sensory changes, or aura-like activity, from stimulation. Last, all experiments were conducted with the subject on their home dosage of anti-epileptic medications to minimize the chances of provoking seizures during research stimulation.

### 2.7 Recording and preprocessing

Neural data from sEEG probes was recorded at a 30 kHz sampling rate with a 1^st^ order high-pass Butterworth filter at 0.3 Hz and a 3^rd^ order low-pass filter at 7.5 kHz on a 256-channel Blackrock Cerebus system (Blackrock Microsystems, UT, USA). Signals were recorded from all sEEG contacts using two contacts visually confirmed to be in white matter as a ground and reference. Custom pipelines were used to process the neural data. For stimulation experiments, a custom artifact removal pipeline was used to remove the stimulation artifact from the neural signals (See Supplementary Material Figure S1). In brief, the pipeline removes 40 samples around each stimulation pulse (10 samples before pulse onset, 30 samples after; 1.34 milliseconds) and replaces their values through linear interpolation. Neural data was then downsampled to 1 kHz and re-referenced using a Laplacian re-referencing scheme (each contact referenced to the average of the two neighboring contacts) to minimize contact specific local common noise. The quality of re-referenced sEEG signals was visually inspected for signs of excessive noise, recording artifact, jaw contraction interference, or excessive interictal epileptic discharges. To ensure an equal number of electrode recordings across experiments, electrode contacts with low-quality signals in any experiment were removed from the analysis of all other experiments as well. Following this preprocessing, 180 electrodes were included in the final analysis.

### 2.8 Data-driven selection of stimulation location

A data-driven selection of the stimulation location was performed by identifying electrode locations displaying spectral differences related to provocative versus neutral trials during the E-GNG task in Exp.1. Spectral decomposition was performed using convolution with a family of complex wavelets with central frequencies ranging from 2-150 Hz (number of cycles ranging from 4 to 12, logarithmically spaced) on the continuous data. Time-frequency power values corresponding to each trial were identified (total time window of 2700 ms: 300 ms before image onset to 400 ms following GNG cue offset, baseline corrected using the first 300 ms before image onset) and compared between provocative and neutral trials using permutation testing (5,000 permutations, alpha value of 0.05, corrected for multiple comparisons using cluster-based correction). The presence of a statistically significant difference in the time-frequency maps for provocative versus neutral trials during the GNG cue presentation (i.e., when the image itself was no longer on the screen) was used as a data-driven approach to guide electrode selection for stimulation experiments.

### 2.9 Analysis of behavior

Accuracy (correct versus incorrect responses) was analyzed using a logistic regression model (RStudio Team [2020] - RStudio: Integrated Development for R. RStudio, PBC, Boston, MA). We reported the change in odds associated with the experimental manipulations as odds ratios (OR) by exponentiating the model coefficients (representing changes in log-odds). First, we assessed the effect of trial type (neutral versus provocative) and of experiment type (Exp.1-4) on overall accuracy. Next, we evaluated the effect of trial type and experiment type specifically on the accuracy of no-go trials. Finally, we analyzed reaction times using all trials where a reaction time was recorded (go trials’ hits, no-go trials’ false alarms) with a general linear model.

Behavioral data collected during the control experiments (standard GNG, subliminal E-GNG) was evaluated for general accuracy and error rates to ensure that the subject was able to perform a standard version of the task, and that a very short image presentation time would reduce, but not cancel, the effect of provocative images. For all experiments (E-GNG, controls) we computed a non-parametric measure of sensitivity (A’; Stanislaw and Todorov, 1999), to determine the ability of the subject to distinguish the go and no-go cues, ranging from 0.5 (chance) to 1 (perfect performance). This measure was computed on each block of 50 trials for each experiment, allowing for a time-resolved visualization of the sensitivity changes across different manipulations and over time.

### 2.10 Analysis of neural data

#### 2.10.1 Changes in spectral power at the stimulation location after neuromodulation

The same analysis carried out to select the stimulation location was performed on that selected location after neuromodulation. This analysis was employed to assess if the biomarker that guided the neuromodulation, i.e. a statistically significant difference in the time-frequency maps for provocative versus neutral trials during the GNG cue presentation of Exp.1 (baseline 1), was still present after neuromodulation (Exp.4, baseline 2). Spectral decomposition and statistical analyses were performed as described above in the section *Data-driven selection of stimulation location.* We performed this analysis on the second baseline only to avoid possible stimulation-related confounds on the spectral content of the signal, ensuring maximal comparability to the analysis performed on the first experiment.

#### 2.10.2 P300 detection: ERP response to GNG cues

Event-related potential (ERP) analysis was accomplished through MATLAB (v2023, MathWorks, MA, USA) using custom scripts. First, the signal from each contact was filtered (1-20 Hz, zero-phase forward and reverse IIR filter) and programmatically evaluated to identify the presence of ERPs occurring during the GNG presentation window (150-450 from cue onset) regardless of the image or signal types (i.e., all trials). The cue window was selected in order to capture the traditional P300 window time (∼300 ms following stimulus onset) as well as common flanking P300 windows seen in deeper neural structures (Fonken et al., 2020; Guex et al., 2020; Linden, 2005).

For P300 detection, we used two criteria (adapted from Bartoli et al., 2019): 1) the amplitude of the ERP in the window of interest was at least 10 times larger than the voltage range detected in a baseline window (500 - 100 ms before image onset) and 2) the standard deviation in the window of interest was at least 2.5 times larger than the standard of deviation in the baseline window. ERP analysis was done for each experiment separately.

#### 2.10.3 P300 modulation: neutral versus provocative trials

Next, electrode contacts displaying a P300 were further evaluated for significant differences in amplitude depending on experimental manipulations. We tested for differences in P300 amplitude between all neutral and provocative trials (overall difference) and for no-go trials only (associated with reactive inhibition). All comparisons between neutral and provocative trials were accomplished via permutation testing with cluster-based correction for multiple comparison (p <0.01). We focused on relative magnitude differences given the variable nature of the polarity of intracranial evoked potentials (as opposed to those measured on the scalp), as that is influenced by the orientation of the electrode with respect to the cortical layer and varies across electrode locations.

#### 2.10.4 Functional connectivity estimation via mutual information

Functional connectivity was estimated through multivariate Gaussian-Copula Mutual Information (shortened here to MI; Ince et al., 2017). All contacts were analyzed for their connectivity to all other contacts (180 electrodes, 16,110 unique pairs), employing pairwise connectivity conditioned on all other recorded signals. For every contact and every E-GNG experiment, the voltage time-series was first filtered into the frequency band of interest (20-60 Hz, based on the results of the time-frequency analysis of Experiment 1) and copula-normalized. We computed the MI during the cue window for each trial (250-500 ms following cue onset) and we evaluated if the exposure to different images affected the functional interactions occurring during the GNG cue window. Overall differences between the MI maps for neutral and provocative trials were assessed using permutation testing (10,000 permutations) for each experiment. Lastly, we evaluated if neuromodulation decreased the context-dependent differences in functional connectivity patterns. First, we identified all pairs of connections with significantly different MI values for neutral and provocative trials (p<0.01), obtaining a binarized set of connections for each experiment (i.e., assigning a value of 1 to connections different for neutral versus provocative trials, 0 for connections not modulated by the image) and we then computed the Jaccard similarity index between the binarized map for Exp.1 and all other experiments.

## 3. Results

Neural data recorded from Experiment 1 revealed the presence of a spectral biomarker in rIFG, which was targeted with neuromodulation in Experiments 2 and 3. Behavioral performance was profoundly affected by exposure to provocative images before neuromodulation, and it improved during and following neuromodulation. After neuromodulation, in Experiment 4, the spectral biomarker in rIFG was no longer present. ERPs and functional connectivity changes were monitored across all experiments. Several recording locations in the brain displayed a P300 in response to the go and no-go cues, and experiments associated with poor performance were characterized by an increased influence of the provocative images over the P300. Lastly, functional connectivity results revealed how brain-wide differences between neutral and provocative trials decreased over the course of neuromodulation, mirroring the behavioral improvement. Each result is reported in detail in the sections below.

### 3.1 rIFG emerges as a neuromodulation target

Combining the data-driven approach with our hypothesis regarding a context-dependent inhibitory control dysfunction, we identified and selected for stimulation an electrode in the rIFG displaying an increased response to the GNG cue during provocative trials (Figure 2A). This biomarker was based on the spectral analysis of recordings obtained during Exp.1, and it constituted an increase in power values following exposure to provocative images, occurring 250-500 ms after the presentation of the go or no-go cue (thus, when there was no image on the screen anymore). This difference occurred across 20-60 Hz. There were other locations displaying differences in time-frequency power values according to the image type, including two adjacent rIFG location, left dorsolateral prefrontal cortex (DLPFC), left putamen, and one location in the left hemispheric white matter (see SI Figure S2). Due to existing evidence on the role of rIFG in action stopping, and the presence of multiple adjacent electrodes displaying the biomarker of interest, we selected rIFG for the neuromodulation experiments (Exp.2-3; Figure 2B).

**Figure 2.**
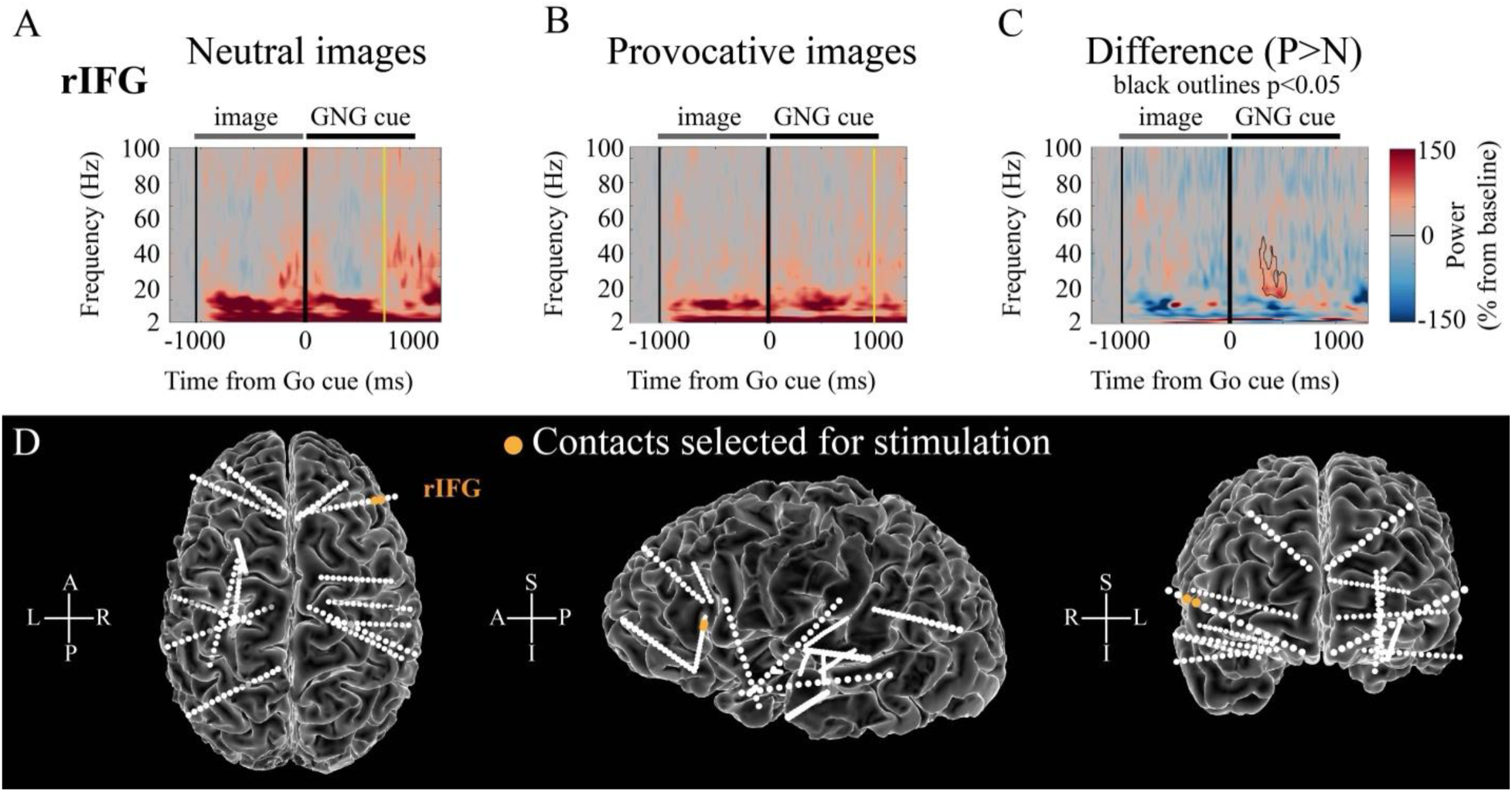
Data-driven selection of stimulation location. A) Activity in rIFG during E-GNG Exp.1 in response to neutral trials, represented as signal power change across the time-frequency space (% change from baseline). Vertical solid black lines mark the onset of the image and of the GNG cue, yellow line shows the median reaction time. B) Same as A for provocative trials. C) Difference between provocative and neutral trials. Areas of statistical significance (permutation testing, p<0.05 corrected) are highlighted by a black outline. An increase in rIFG power for provocative trials occurred after about 250 ms from the cue presentation, between 20-60 Hz. The difference occurred before the median RT of either neutral or provocative trials. D) Sagittal, coronal, and axial projections of the cortical surface showcasing sEEG placement. Electrode contacts that were selected for stimulation in Exp.2 and 3 (rIFG) are highlighted in orange. The selection was driven by the results shown in panel C.

### 3.2 Task performance is affected by provocative images

Overall accuracy in the E-GNG task was significantly different between neutral and provocative trials, with the subject performing significantly worse when exposed to provocative stimuli (logistic regression model, provocative versus neutral: OR = 0.109, p<0.001). Across all experiments and regardless of go/no-go round status, neutral trial accuracy remained high and relatively unchanging, ranging between 95% and 99% accurate. Provocative trial accuracy varied significantly, ranging from 66% to 93% accurate (Table S1 and Figure 3A). The different experiment types affected the odds of a correct response when compared to the first E-GNG baseline (Exp.1), and we repeated the logistic model analysis separately for the neutral and provocative data in order to better isolate the effects of stimulation.

**Figure 3.**
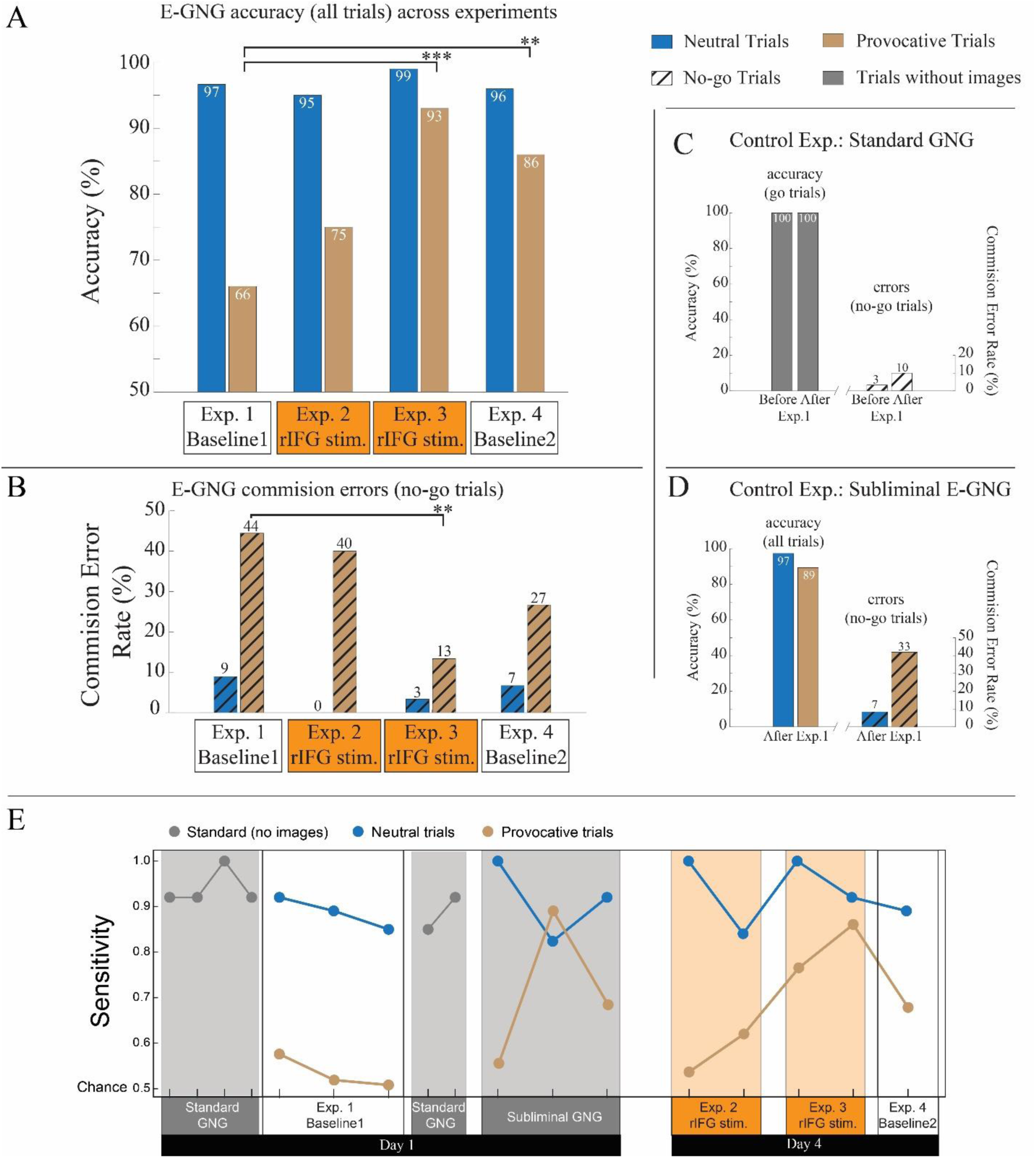
E-GNG performance. A) E-GNG overall accuracy (all trials), divided by exposure to neutral or provocative stimuli for each experiment. The panel shows the modulation in performance across experiments, starting from the first baseline (Exp.1), stimulation of rIFG during image presentation (Exp.2), continuous stimulation of the rIFG (Exp.3), and second baseline (Exp.4). During Exp.1, overall accuracy was significantly worse following provocative image exposure when compared to neutral trials. Accuracy improved with stimulation, reaching its maximum during continuous stimulation of the rIFG (Exp.3). Accuracy then decreased (baseline 2, Exp.4) but was still significantly higher than overall provocative accuracy for Exp.1. Asterisk denote significant differences based on the OR model results (** p<0.01, *** p<0.001). B) Same data as in A, but showing only no-go trial commission errors (incorrect responses to no-go trials). The pattern of improvement specifically during continuous rIFG stimulation (Exp.3) is even more evident for commission errors. C) Go accuracy and commission errors for the standard GNG control task. The two bars correspond to the standard GNG task performed before Exp.1 and the standard GNG performed right after Exp.1. The patient showed perfect performance on go trials and low commission errors, in line with values obtained for neutral trials in the first E-GNG experiment. D) Overall accuracy and commission errors for the subliminal E-GNG control, performed after Exp.1. The overall performance shows some modulation for provocative trials, and the evaluation of the no-go trials demonstrates a high rate of commission errors in the provocative condition driving the difference. E) Non-parametric sensitivity analysis of subject’s ability to distinguish go vs no-go cues across controls and experiments (A’, 0.5 = chance, 1 = perfect performance). Sensitivity values are computed on each block of 50 trials to have a time-resolved measure of sensitivity changes also within experiments. Each marker represents the value for a block of 50 trials, color-coded by experiment and condition.

Exp.1 (baseline 1) had the lowest overall provocative accuracy of 66% (go rounds 70%; no-go rounds 56%). Using Exp.1 as the logistic model baseline, the odds of success were significantly higher during continuous stimulation of rIFG, leading to 93% accuracy (Exp.3: OR = 6.84, p < 0.001). This significant increase in odds was maintained in the absence of stimulation during the second baseline (Exp.4: OR = 3.17, p = 0.009), although with overall accuracy values lower than continuous stimulation of rIFG (86%). The only experiment not showing a significant provocative trial performance improvement was the rIFG image-locked stimulation, with 75% accuracy (Exp.2: OR = 1.55, p = 0.131). Thus, with respect to the first E-GNG task performed by the patient (Exp.1), provocative trial accuracy improved significantly with continuous stimulation of the rIFG, with stimulation effects carrying over in the absence of stimulation (Exp.4).

To more directly assess inhibitory control changes, neutral and provocative models were adjusted to analyze the influence of neuromodulation on no-go trial accuracy (Figure 3B). Similar to overall accuracy, the accuracy of no-go trials preceded by neutral images was unaffected by any form of stimulation (all p>0.05): commission errors were low at baseline (9%) and throughout all experiments (between 0% and 9%). On the contrary, provocative no-go trial commission errors were high at baseline (Exp.1: 44%) with the only significant decrease occurring with continuous stimulation of the rIFG (Exp.3: no-go accuracy improvement, OR=5.2, p=0.007), reaching a low of 13%. Image locked stimulation did not result in a significant improvement in no-go performance with respect to the first baseline.

In summary, no-go accuracy was most impacted by exposure to provocative images, and it was significantly improved only by continuous stimulation of rIFG (Exp.3). Overall accuracy (including both go and no-go trials) was also affected by presence of provocative images, although a significant improvement was more robustly obtained across multiple experiments (Exp.3 and 4). Lastly, reaction time of correct go trials analysis demonstrated there was no significant difference in the overall speed of response when exposed to provocative versus neutral stimuli (average and standard deviation: 581.9±229.4 ms; versus 570.4±77.3 ms, beta = 3.97, p=0.79), although we note that the variation in the reaction times was more than doubled in the provocative trials.

### 3.3 Control experiments

#### 3.3.1 Standard GNG

The standard GNG task was performed directly before and after the first E-GNG experiment (Exp.1) to control that the patient was not experiencing non-specific disruptions in his cognitive or attentive abilities that would affect interpretation of the E-GNG results. The participant performed well on the standard GNG task, with 100% accuracy on go cues both before and after the first E-GNG experiment (before: 140/140 correct go cues, after: 70/70 correct go cues; Figure 3C). Commission errors (responses to no-go cues, false alarms) were low before (3/60 no-go responses, 5%) and somewhat increased after the E-GNG (3/30 no-go responses, 10%), although this difference was limited in absolute terms (3 errors).

#### 3.3.2 Subliminal E-GNG

As a final control, we analyzed behavior in response to a subliminal version of the E-GNG, in which the images (both neutral and provocative) were briefly flashed for 50 ms (rather than presented for 1000 ms). This control was performed to evaluate if even a very brief, almost subliminal presentation of the images was still capable of inducing a specific disruption of the E-GNG performance. Overall accuracy showed somewhat lower values for provocative trials (neutral: 97%; provocative: 89%; Figure 3D). After subdividing for go cues and no-go cues, it was clear that the accuracy for go cues was high and identical between neutral and provocative trials (104/105 for both, 99%) while the accuracy for no-go cues was low, with increased commission errors for provocative trials (15/45, 33.3%) but not for neutral (3/45, 6.7%; Figure 3D). These results indicate that provocative images specifically affected inhibitory control abilities (no-go trials) when presented subliminally (logistic regression model, change in odds of correctly inhibiting the response to no-go cues for provocative versus neutral, OR = 0.14, p=0.004).

### 3.4 Sensitivity changes within and across all experiments

Sensitivity (A’) was close to 1 for standard GNG trials (ranging between 0.85-1) and for neutral block of trials during the E-GNG experiments (0.82-1; Figure 3E). Contrarily, sensitivity was low during provocative trials in the first E-GNG experiment (0.51-0.57). Some within-experiment variability in sensitivity is visible for the subliminal provocative trials during the control E-GNG experiment, reaching quite high values in the middle of the task (A’=0.89 in the second block of 50 trials of subliminal E-GNG). Sensitivity increased steadily with neuromodulation in Exp.2 (from 0.53 to 0.61) and had a big improvement with Exp.3, starting at 0.53 and arriving at 0.86. After stimulation was turned off in Exp.4, a decrease in sensitivity can be observed (A’=0.67), but not to the level of Exp.1 and 2, indicating some carryover effect of continuous stimulation that was maintained. The observation that sensitivity was reduced specifically following provocative images, and that systematic improvement in sensitivity occurred only during stimulation experiments, confirms that the improvement in performance cannot be ascribed to repetition effects, as those should also occur in experiments without stimulation.

### 3.5 rIFG biomarker is no longer detected after neuromodulation

We repeated the time-frequency analysis on the electrode in the rIFG selected for neuromodulation after stimulation (Exp.4, baseline 2). The biomarker (increased 20-60 Hz response to the GNG cue during provocative trials) was not present following neuromodulation (all p>0.05 in the time-frequency difference analysis, Figure 4A). Additionally, all other locations that displayed a significant difference between provocative and neutral trials in Exp.1 (Figure S2) were no longer statistically different in Exp.4 (Figure S3). This further validates that the influence of provocative images over the GNG cue processing, assessed with the time-frequency analysis, was reduced or even eliminated following neuromodulation.

**Figure 4.**
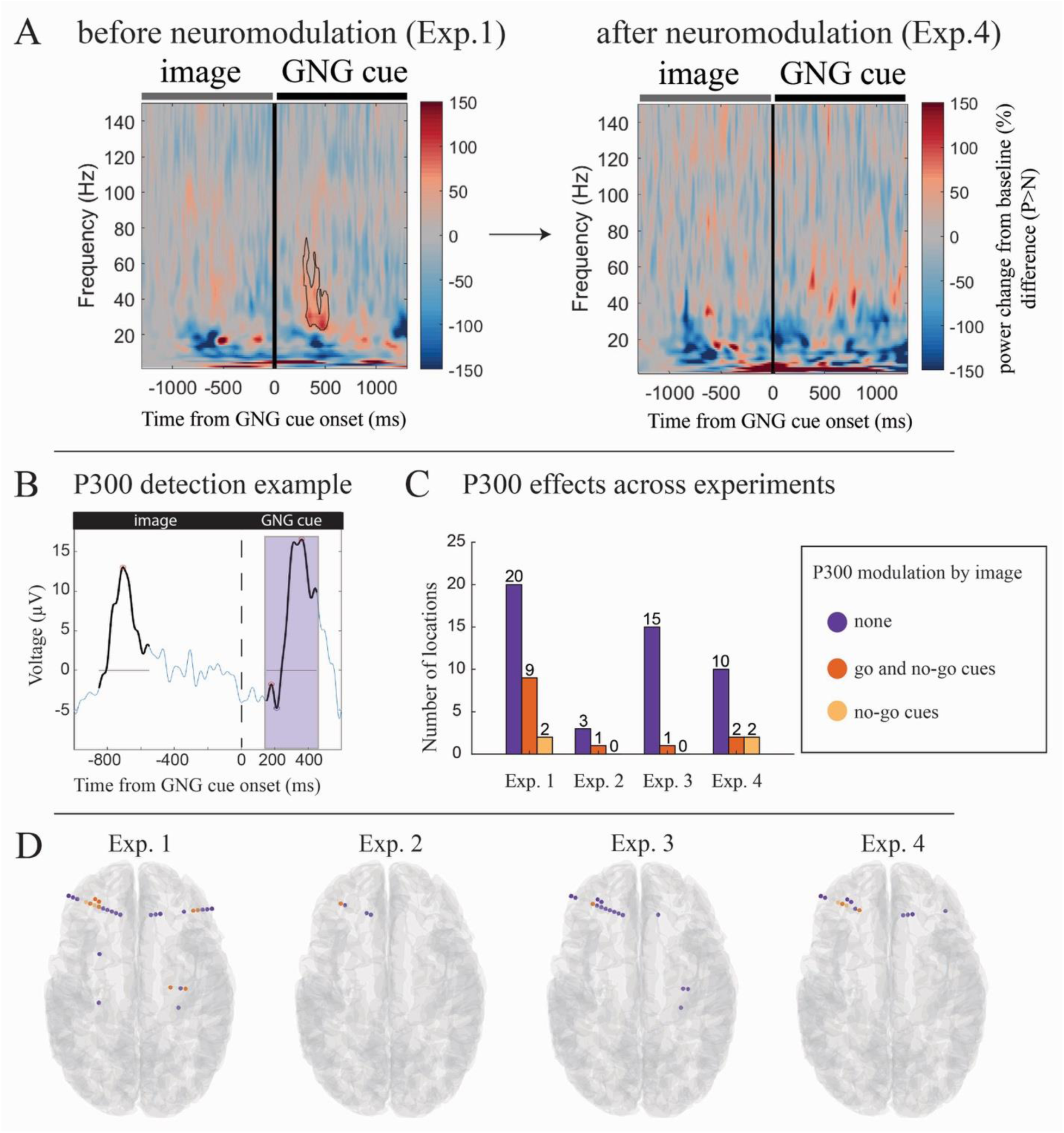
Spectral and P300 changes across experiments. A) Time-frequency analysis of the rIFG before (Exp.1, same data as shown in Figure 2. C) and after (Exp.4) neuromodulation. The significantly elevated activity in the cue window at 20-60 Hz in Exp.1 no longer present in Exp.4. B) P300 detection example in one electrode location, showing voltage values averaged across all trials in blue, and values exceeding the threshold for detection marked in black. ERPs in response to image presentation are not considered. ERPs occurring between 150-450 ms from the GNG cue onset (purple shading) are classified as P300. C) Bar graph displaying the number of electrodes displaying a P300 in response to the GNG cues, separated into three categories: P300 with no amplitude modulation by image type, P300 modulated by image type (considering both go and no-go cues, p<0.01), P300 modulated by image type in no-go trials only (p<0.01). D) Electrode locations displaying a P300 are superimposed onto the subject’s brain and color-coded according to the presence of modulation by image type.

### 3.6 GNG P300 modulation by image type is associated with poor performance

We detected locations exhibiting a P300 to the GNG cues (Figure 4B). We then tested whether those locations were exclusively responsive to the cues, or whether they showed additional modulations according to the previously presented image. Exp.1 had 29 contacts displaying significant P300 activity in response to the GNG cues, 31% of which (n=9) additionally displayed a significant modulation depending on the image type (amplitude difference p<0.01 with cluster-based correction across the time window). Thus, the presentation of the provocative images affected the neural response to the following GNG cue.

We monitored the P300 activity in those 29 contacts across the stimulation experiments and second baseline to evaluate changes across experiments (Figure 4C and 4D; see S4 for additional information). During image-locked stimulation of the rIFG (Exp.2) 4 of the locations identified in Exp.1 showed a detectable P300, with 25% of those (n=1) displaying a significant modulation depending on the image. During continuous stimulation of rIFG, 16 contacts displayed a P300, and only 6.3% (n=1) were modulated by the images. Lastly, during the second baseline (Exp.4), we detected a P300 on 12 contacts, with 16.7 % (n=2) being modulated by the image type. Next, we restricted the analysis to the no-go trials only, and only in Exp.1 and 4 we could detect contacts displaying a P300 in response to the no-go cues that were significantly modulated by the images. All detected contacts were located in the left frontal cortex (n=2 in Exp.1, same locations that also displayed a P300 modulation when considering all trials; n=1 in Exp.4, specific to no-go trials only; see Table S2). Overall, behavioral improvements across experiments were mirrored by a reduction in the proportion of locations displaying a P300 that was modulated by the provocative images (lowest for Exp.3).

We refrain from interpreting the anatomical distribution of the effects in detail due to concerns of structural anomalies related to the left temporal resection. For reporting purposes, we note that the P300 responses were detected across bilateral locations in IFG, orbitofrontal cortex and medial temporal cortex, in the left DLPFC and putamen, and in the right hippocampus (Figure 4D).

### 3.7 Functional connectivity differences are reduced after neuromodulation

We evaluated if provocative and neutral images affected functional connectivity patterns estimated during the presentation of the GNG cue (Figure 5A). Permutation testing of the MI maps for neutral and provocative trials revealed there was a significant overall difference in the MI values for both Exp.1 and 2 (p<0.0001 for both experiments), while Exp.3 and 4 showed no overall significant differences (p = 0.109; and p=0.907). Thus, experiments associated with poor GNG performance on provocative trials displayed a significantly different set of functional connections during the GNG presentation for neutral and provocative trials. This difference was no longer significant during and after continuous rIFG stimulation, mirroring the behavioral improvement in these experiments.

**Figure 5.**
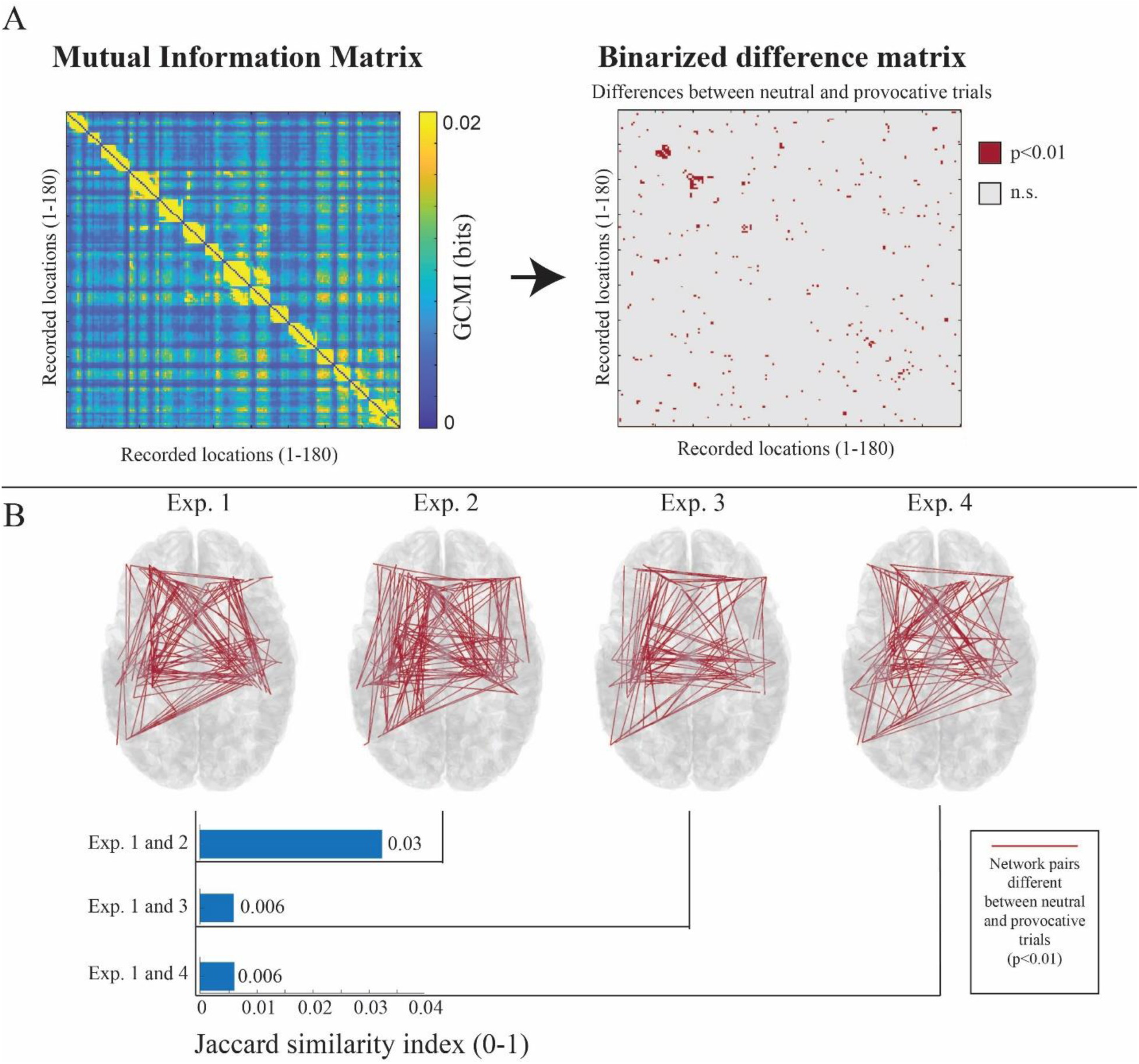
Mutual information difference matrix and dissimilar functional connectivity maps. A Example of the creation of the difference matrix. For each experiment we estimated the functional connectivity values using mutual information (MI) between each pair of contacts (recording locations) for each trial. These maps were then binarized based on the presence of significant differences between neutral and provocative MI values (p<0.01). B) Statistically significant differences (p<0.01) in functional connectivity between neutral and provocative trials, superimposed over the subject’s brain for visualization purposes. The number of pairs with functional connectivity differences between neutral and provocative trials is higher and more similar (based on the Jaccard similarity index) in experiments with poorer behavior outcomes (Exp.1 and 2) versus the experiments with significantly improved behavior (Exp.3 and 4).

Next, we assessed the changes between experiments more directly. We tested the MI values difference between neutral and provocative trials for each connection pair in each experiment (p<0.01) and obtained a binarized network graph capturing the connections modulated by the image type. Exp.1 and 2 displayed a large number of electrode pairs with significant differences in connectivity depending on stimuli exposure type (205 and 240 significant differences, respectively), while Exp.3 and 4 had about half as many electrode pairings with significant differences in their connectivity based on trial image type (131 and 124, respectively). Additionally, the graph for Experiment 1 revealed higher rates of similarity to Exp.2 (Jaccard index = 0.03) than to Exp.3 or 4 (Jaccard index = 0.006 for both), indicating that the functional connectivity differences for neutral and provocative trials were more pronounced and spatially similar in the first baseline and during time-locked rIFG stimulation, which are the two experiments associated with poor behavioral performance to provocative stimuli. With continuous rIFG stimulation and in the following baseline, we found a reduction in the number of connections sensitive to the image presentation and a decrease in network similarity with respect to the first baseline, in line with the improved behavior, no longer affected by the provocative images (Figure 5B).

## 4. Discussion

The goal of this single-subject study was to test whether the patient’s inhibitory control processes were affected by stimuli related to his KBS pathology, evaluated with a personalized emotional GNG paradigm tailored to provoke the hypersexual symptoms of the patient (Figure 1). Furthermore, this study aimed to determine whether a detectable biomarker of the patient’s symptoms was present in their neural data, and whether personalized, targeted neuromodulation could lead to trackable changes in this biomarker as well as an improvement in the subject’s behavior.

In line with our hypothesis, the patient demonstrated poor action suppression/inhibition abilities when presented with provocative visual content. Excessive attentiveness to visual stimuli, also known as hypermetamorphosis, is a known symptom of KBS (Willis and Haines, 2018), and likely contributing to exacerbating the patient’s symptoms during provocative image exposure. A clear biomarker was identified in the rIFG, where signals from multiple electrodes displayed increased gamma range activity to the presentation of the go and no-go cues following provocative images, but not neutral images (Figure 2). This increased response constitutes evidence that the presentation of provocative images altered the subsequent processing of the go and no-go cues, likely leading to the poor performance in the task (Figure 3). This effect was also visible in the ERPs, as several locations generating a P300 in response to the go/no-go cues were also influenced by whether the image preceding the cue was neutral or provocative (Figure 4). Data-driven neuromodulation of the rIFG was effective at improving the behavioral performance of the patient, and we evaluated associated changes in neural activity during and after neuromodulation.

### 4.1 Provocative images affect inhibitory control abilities

The subject’s overall accuracy was significantly affected when exposed to provocative stimuli during the E-GNG paradigm. Such results are particularly significant given that, in a similar paradigm, the accuracy of healthy controls was not modulated by the emotional content of the images paired with the GNG task. Normally, more sensitive paradigms and measures of behavior are needed to highlight the interaction between image content and behavioral performance, such as the stop-signal reaction time (Littman and Takács, 2017a).

The subject’s performance in the control experiments (standard GNG and subliminal E-GNG) highlights the specificity of this finding. First, the patient was able to perform a standard GNG paradigm with high accuracy, providing further support that the patient’s action-stopping abilities were not generally impaired, but the effect was only revealed in the presence of emotional triggers. Critically, the patient displayed perfect accuracy to go-cues in the standard control, ensuring that there were no attentive or cognitive issues affecting his performance, and confirming that the behavioral deficits observed in the first E-GNG task could not be explained by some confounding variable other than the presence of the provocative images. Second, during the subliminal version of the paradigm, we found high accuracy scores for both neutral and provocative trials in go trials, but low provocative accuracy in no-go trials. This result seems to indicate that even brief exposure to provocative stimuli can have a significant effect on the subject’s impulse control abilities, in line with existing evidence that subliminal stimuli can have a significant influence on inhibitory control (Parkinson and Haggard, 2014). With this control experiment, we could clearly identify a specific deficit in inhibitory control (no-go trials). The fact that the patient could perform a standard version with no issue and that the subliminal presentation isolated the effect on no-go trials, altogether support our hypothesis of a context-specific inhibitory control issue that is triggered by provocative stimuli.

### 4.2 Neuromodulation to the rIFG rescues performance

Neuromodulation improved accuracy across provocative trials, with continuous stimulation of the rIFG generating the largest improvement. Neutral trial accuracy was high at baseline and remained unaffected by any form of tested neuromodulation or repetition of experimental material. Provocative trial accuracy improved significantly with neuromodulation, particularly with continuous stimulation of the rIFG, which seemed to have a lasting effect. Indeed, turning off the stimulation caused a decrease in the performance, but not to the same level as before stimulation. When considering commission errors alone (responses to no-go trials), only continuous rIFG stimulation caused a significant improvement with respect to the first experiment, demonstrating the specificity of rIFG stimulation in modulating action-stopping abilities (Wessel et al., 2013b) and the need of sustained stimulation to reach a significant effect. This supports the idea that the rIFG is a key player in at least response inhibition behavior, if not general inhibition (Aron et al., 2014; Hampshire et al., 2010; Littman and Takács, 2017a). Furthermore, the sensitivity analysis showcased a reliable cumulative improvement across blocks of provocative trials only during rIFG stimulation, with the steepest increase during continuous stimulation, dissociating the changes from repetition effects, which should occur within each experiment.

The lopsided nature of the neutral versus provocative accuracy results indicates that the inhibitory control issues become evident once the subject is exposed to some form of sexually provocative cue. This demonstrates that the patient does not exhibit a general inhibitory control issue, and that the exposure to provocative images could be interfering with the ability to engage the inhibitory control circuit appropriately. Rather than purely inhibitory in origin (Parvizi, 2012), this could be related to a biased weighting of the value of action-stopping in a provocative context, where the reward of engaging with the images overrides ongoing task goals. This outcome is also congruent with the theorized hypersexuality etiology of a preexisting pathology being present in the subject’s remaining intact temporal lobe, something that was previously controlled by the now resected left temporal lobe (Baird et al., 2002). This is further supported by the significant improvement that continuous neuromodulation has on behavior. Similar to how DBS regulates the occurrence of aberrant communication in neuropsychiatric conditions (Horn et al., 2019; Neumann et al., 2023; Provenza et al., 2024; Reddy et al., 2024), the continuous stimulation of the rIFG could be engaging the action-stopping circuit notwithstanding the effect of the provocative images, thus bringing the subject’s neural activity closer to a healthy state.

### 4.3 Data-driven selection of the stimulation location based on a spectral biomarker

A goal of this study was to identify and monitor a neural biomarker associated with the abnormal behavior demonstrated by the subject. To this end, we evaluated the spectral content of intracranially measured signals across a wide time-frequency space during the E-GNG task, searching for locations exhibiting increased activity specifically for provocative trials. Significantly heightened activity in the rIFG in response to provocative stimuli during the cue presentation in Exp.1 was identified as a potential biomarker during this study due to its location and its window of occurrence. The rIFG has been found to be a key contributor to most, if not all, kinds of action-stopping processes (Aron, 2011), including the suppression of unwanted thoughts (Apšvalka et al., 2022). Furthermore, its activity being significantly different during the cue presentation window (i.e., go or no-go cue), when images are no longer visible, corresponds with our hypothesis that provocative exposure is exerting a prolonged influence, bleeding beyond the visual processing stage and influencing the ability of the rIFG to initiate action-stopping. Indeed, recent evidence indicates that patients with rIFG lesions have difficulties in initiating, rather than implementing, action-stopping (Choo et al., 2022).

Consequently, we tested whether we could mitigate or remove this influence with rIFG direct stimulation, and if the subject’s behavior could be rescued. Following neuromodulation, in Exp.4, the time-frequency based biomarker in rIFG was no longer present, and the behavioral performance of the patient was significantly improved with respect to the first experiment, as discussed above. This could suggest that the continuous stimulation of the rIFG in Exp.3 has had an influence on its activity even after stimulation ceased, something that has been demonstrated previously in time-frequency analysis studies (Bosch et al., 2025; Ghin et al., 2021). This phenomenon would not be entirely unexpected and matches current literature surrounding the immediate and lasting impact of stimulation on aberrant neural circuits (Harmsen et al., 2022; Neumann et al., 2023; Provenza et al., 2024; Reddy et al., 2024).

Noteworthy, the same pattern (i.e., no statistically significant differences in the time-frequency power values after neuromodulation) was observed across all other locations in the brain that displayed significantly heightened activity upon exposure to provocative stimuli in Exp.1 (Figures S2 and S3). These locations spanned areas involved with cognitive and inhibitory control, such as the dorsolateral prefrontal cortex (Gavazzi et al., 2023) and putamen (Guo et al., 2018), plus a white matter location. This finding suggests that the effect of stimulation was not constrained to the rIFG alone, but it influenced other functionally interconnected regions, in a way that was sustained also after stimulation was turned off.

### 4.4 Changes in P300 before, during and after neuromodulation

The spectral analyses were used to guide neuromodulation, by selecting a location based on the presence of a biomarker (i.e., increased activity for GNG cues following provocative versus neutral trials) and to verify if the biomarker was changed following neuromodulation. We additionally quantified neural activity before, during and after neuromodulation using ERPs, focusing on the P300, a component occurring between 250-400 ms after the presentation of go/no-go cues and displaying a positive polarity in scalp EEG studies.

The P300, unlike other recognized ERPs such as the N2 or the P300’s subsets like P3a and P3b, was chosen as it is reliably found throughout several regions such as the hippocampus, amygdala, prefrontal cortex, and anterior cingulate cortex in response to go and no-go cues as well as salient stimuli (Alho et al., 2015; Diesburg et al., 2024; Fonken et al., 2020; Huster et al., 2020; Linden, 2005; Matsuda et al., 2023). The influence different types of stimuli have on the P300 during a GNG paradigm has been studied repeatedly with scalp EEG (Albert et al., 2010; Zhao et al., 2019), as well as in other traditional well established paradigms such as the oddball task and the stop-signal task (Camfield et al., 2018; Zhao et al., 2024). In contrast to our study, most paradigms combining GNG with emotional images tend to use the emotional stimuli (images or words) as the go and no-go cues, thus conflating the image (or word) processing with the action-suppression cue (Chiu et al., 2008; Wang et al., 2011; Yang et al., 2014). While the direction of the effects is somewhat variable among these studies, perhaps due to the different stimuli and paradigms, they converge on the presence of a modulation of the P300 for emotional versus neutral stimuli. Thus, the effect of emotional stimuli that do not overlap with the go and no-go cues is less explored. Here, we demonstrate that task-irrelevant provocative stimuli, preceding the presentation of the go and no-go cues, were able to influence the P300 in response to the cues. Several locations exhibited a P300, and a large proportion of these (31%) were modulated by the provocative versus neutral images in the first experiment. While the number of locations displaying a P300 varied across experiments, the proportion of the P300 sensitive to the provocative images showed a clear pattern, decreasing during image-locked rIFG stimulation (25% in Exp.2) reaching its minimum during continuous stimulation (6% in Exp.3) and somewhat increasing again during the last experiment in the absence of neuromodulation (17%, Exp.4). This pattern mirrors the changes observed in behavioral measures, with the worst performance associated with the highest proportion of locations exhibiting a P300 modulated by the provocative images, and the best performance, especially in commission errors, being paired with the least amount of P300 modulations.

These results seem to indicate that the P300 in response to the go and no-go cues needs to be relatively immune to any sort of contextual factors (i.e., previously seen image) in order to lead to successful action-stopping capabilities. This aligns with evidence from non-invasive studies, as the effect of emotional and arousing stimuli on ERP activity often translates into higher rates of inhibition errors or slower reaction times (Fisher et al., 2011; Gaspelin et al., 2023; Hu et al., 2023; Johnstone et al., 2013; Xu et al., 2015).

### 4.5 Wide-spread connectivity changes reflect improvement

Finally, we estimated mutual information as a proxy of functional connectivity during the time window and frequency range defined by the rIFG biomarker (250-500 ms after GNG cue onset, 20-60 Hz). Existing non-invasive studies assessing functional connectivity in relation to an emotional variant of the GNG paradigm focus on determining which region is active at specific points throughout the paradigm, and with which other regions communication is flowing, rather than focusing on the relationship between connectivity and behavior (Lapate et al., 2022; Lee et al., 2018; Stoycos et al., 2017). In contrast, our goal was to investigate the neural activity behind the different behavior exhibited in response to the GNG cues. Specifically, we aimed at identifying differences in brain-wide connectivity following the exposure to neutral and provocative images, and tracking those differences during and after rIFG neuromodulation. The behavioral difference between neutral and provocative trials in the first two experiments was reflected in the overall functional connectivity, as the MI-based networks for neutral and provocative trials were significantly different. This overall difference is compatible with a wide-spread effect of provocative image exposure in the patient, introducing sustained differences in interareal information exchange also after the image presentation, potentially underlying the failure to correctly react to the cue. During and after continuous rIFG stimulation the difference in the overall functional connectivity patterns was reduced to the point that it did not reach statistical significance, matching the observed improvement in behavioral performance. Indeed, several brain regions spanning parietal, cingulate and frontal cortex are modulated not only by the presence of emotional images in go/no-go task, but also by their task-relevance (Mancini et al., 2022; Mirabella et al., 2024). While the emotional images in our paradigm were all task-irrelevant (i.e., they simply preceded the presentation of the go/no-go cue)(Littman and Takács, 2017b), it is likely that the subjective relevance of the provocative images for the patient was initially high, and then reduced with neuromodulation leading to the observed non-significant differences in interareal communication with respect to the neutral images.

The network similarity results further confirmed that the experiments with poorer behavior outcomes (Exp.1 and 2) did not only have a higher number of functional connectivity differences between neutral and provocative trials, but that these differences were highly preserved across both experiments in terms of spatial distribution across the brain (Figure 5). Indeed, the similarity between the first two experiments is six-fold the similarity between Exp.1 and the later experiments (Exp.3 and 4), which in turn are associated with an increased overlap in functional connectivity between neutral and provocative trials, and improved behavioral performance for provocative trials. The unique effect that provocative images had on the patient was therefore detectable also in the whole-brain functional connectivity patterns, and changes in the connectivity mirrored changes in task performance during and after neuromodulation. Continuous, high-frequency stimulation of the rIFG may have been able to override the aberrant response to provocative images, bringing the network activity closer to a neutral state. Coupling neuromodulation with cognitive interventions (Soleimani et al., 2025; Spagnolo et al., 2020) is a novel avenue to maximize stimulation-induced plasticity (Kricheldorff et al., 2022). Altogether, this further supports the idea that the targeted neuromodulation is preventing the underlying pathology from manifesting symptomatically.

### 4.6 Limitations

There are a few limitations with the current study. Like all case studies, it is difficult to generalize findings or compare results against healthy controls, an issue that is particularly exacerbated here as the limited number of other studied cases and the various potential etiologies of those cases make it difficult to even compare against others displaying similar symptoms. To mitigate these issues, we performed several control experiments, and we related our behavioral and ERP findings from healthy controls performing similar paradigms.

The number of experiments and experimental manipulations was constrained by clinical factors. For example, it is impossible for us to disentangle whether the strong behavioral improvement obtained during the continuous stimulation of rIFG (Exp.3) could also have been obtained with a prolonged version of the image-locked stimulation (Exp.2). This presence of an additive effect would not be unexpected as targeted stimulation effects on cognitive symptoms show improvement over time with continuous treatment (Ranjan et al., 2024; Sheth and Mayberg, 2023). Similarly, it is difficult to predict for how long the lingering positive influence of continuous stimulation should be expected to last, especially as the neuromodulation session was quite brief and typically weeks are required for a therapeutic response to build up.

Nonetheless, our work demonstrates the possibility of creating targeted, data-driven personalized neuromodulation and obtaining real-time improvements in patient performance. Future studies will be needed to generalize this approach for those suffering from other inhibitory control deficits or executive dysfunction. Analyzing the long-term influence of neuromodulation on subjects experiencing hypersexuality, as currently done for those with depression and OCD, could determine if neuromodulation is a viable potential therapy for those with the condition who have failed other treatments. Finally, other case studies could evaluate the viability of the process used in this case study, data-driven neuromodulation targeting of specific neural regions based on responses to a well-established paradigm, to ascertain the potential of this method as a form of personalized medical care (Soleimani et al., 2025).

## 5. Conclusions

Our study indicates that the patient’s KBS symptoms cannot be ascribed to a general lack of inhibition, in line with the argument that “disinhibition” is an over simplification of the cognitive processes underlying failures in action-restraint (Parvizi, 2012). Indeed, the patient displayed intact inhibitory abilities in standard and neutral contexts, while abnormal behavior occurred systematically once exposed to provocative material. We leveraged the unique opportunity to investigate the neural activity underlying the patient’s behavior, and delivered data-driven personalized neuromodulation based on a custom-biomarker. Neuromodulation rescued the inhibitory control performance to the level obtained in neutral contexts, and it also reduced all neural signatures associated with abnormal behavior, spanning regional spectral features, event-related potentials and functional connectivity. We can only speculate on what could be the effect of neuromodulation on the underlying cognitive processes. Continuous stimulation to the rIFG could have lessened the attentional capture effect of provocative stimuli (Camfield et al., 2018; Hajcak et al., 2010), supported by evidence suggesting that rIFG plays a key role in detecting salient events and pausing ongoing processes (Choo et al., 2022; Corbetta et al., 2008; Diesburg and Wessel, 2021a, 2021b; Erika-Florence et al., 2014). Alternatively, stimulation could have increased the value associated with action-suppression, increasing proactive control (Cai et al., 2017b, 2011; Chikazoe et al., 2009; Swann et al., 2012) and eventually prioritizing the current goals over engaging with provocative content. Our work’s main outcome is the proof of concept for the approach, rather than an immediate therapeutic option for the patient, especially given that the rIFG is not an approved neuromodulation target and it would require a clinical trial to be employed as such. From a practical standpoint, the patient may benefit from cognitive-based therapeutic strategies incorporating conditioning elements with exposure therapy.

## Data Availability

All data produced in the present study are available upon reasonable request to the authors

## Acknowledgments

We thank the patient and his family for taking part in this personalized research protocol. We are grateful to the hospital staff for their assistance in keeping the patient comfortable and cared for throughout the study.

## CRediT

**Layth Mattar** - data curation, formal analysis, software, validation, visualization, writing - original draft

**Shraddha Shah** - conceptualization, investigation

**Lily S. Chamakura** - resources, software

**Denise Oswalt** - resources, software

**Yue Zhang** - resources, investigation

**Davin Devara –** investigation, writing - review and editing

**Jung Uk Kang** - writing - review and editing, visualization

**Zahra Jourahmad** - writing - review and editing

**Ryan Jafri** - writing - review and editing

**Geoffrey Liu** - writing - review and editing, data curation

**Joshua Adkinson** - investigation, software

**Isabel A. Danstrom** – investigation

**Xiaoxu Fan** - resources, investigation

**Yvonne Reed** – investigation, writing - review and editing

**Kelly R. Bijanki** - supervision, writing - review and editing

**Alica Goldman** - supervision, writing - review and editing

**Lu Lin** - supervision, writing - review and editing

**Vaishnav Krishnan** - supervision, writing - review and editing

**Nicole R. Provenza** - writing - review and editing

**Andrew J. Watrous** - investigation, writing - review and editing

**Sarah R. Heilbronner** - project administration, supervision writing - review and editing

**Sameer A. Sheth** - funding acquisition, project administration, writing - review and editing

**Garrett P. Banks** - conceptualization, project administration, investigation, supervision, writing - review and editing

**Eleonora Bartoli** - conceptualization, methodology, investigation, supervision, software, writing - review and editing

## Supplementary Material

**Table S1.** Behavioral accuracy of Exp.1-4, subdivided into neutral and provocative trials and go and no-go trials.

|  | Experiment | Total Number of Trials | Accuracy Neutral Trials (%) | Accuracy Provocative Trials (%) | Accuracy Go Trials (%) |  | Accuracy No-go Trials (%) |  |
| --- | --- | --- | --- | --- | --- | --- | --- | --- |
|  |  |  |  |  | N | P | N | P |
| Exp.1 | No Stimulation | 300 | 97 | **66** | 99 | 70 | 91 | **56** |
| Exp.2 | rIFG stimulation locked to image | 200 | 95 | 75 | 93 | 81 | 100 | 60 |
| Exp.3 | rIFG continuous stimulation | 200 | 99 | **93** | 100 | 96 | 97 | **87** |
| Exp.4 | No Stimulation | 100 | 96 | 86 | 97 | 91 | 93 | 73 |

**Table S2.** Number of electrode contacts displaying a P300 in response to the go or no-go cues and their modulation by image type. The table summarizes the number of contacts that display a P300 (first column) and then subdivides depending on whether they displayed a significant modulation of the P300 amplitude by the image presented before the GNG cue. Experiment 3, associated with the best performance in the task, displayed the lowest proportion of contacts with a P300 to the GNG affected by the image type.

| Experiment | Contacts with a P300 in response to the GNG Cue |  |  |  |
| --- | --- | --- | --- | --- |
|  | Total Contacts | Number of contacts with a P300 not modulated by image type | Number (and %) of contacts with a P300 modulated by image type (N vs P difference) | Number of contacts with a P300 modulated by image type (N vs P trials in no-go trials) |
| Exp.1 | 29 | 20 | 9 (31%) | 2 |
| Exp.2 | 4 | 3 | 1 (25%) | - |
| **Exp.3** | **16** | **15** | **1 (6.3%)** | **-** |
| Exp.4 | 12 | 10 | 2 (16.7%) | 2 |

### Stimulation artifact rejection

The artifact rejection pipeline employed for neural recordings obtained during Experiments 2 and 3 is depicted in Figure S1. First, we identified the onset of each stimulation pulse (within the train of pulses) and blanked 10 samples before and 30 samples after the pulse onset for the signals recorded from all electrodes. Linear interpolation and shift adjustment were then applied to avoid sharp transitions between the signal edges. This method created a loss of data of 40 samples (1.33 ms for recordings performed at 30kHz sampling rate, like in our case). Importantly, the same time window is blanked across all recording electrodes, thus not generating any biases in mutual information metrics (i.e., mutual information is identical across all recordings for those samples, not contributing to any differences).

**Figure S1.**
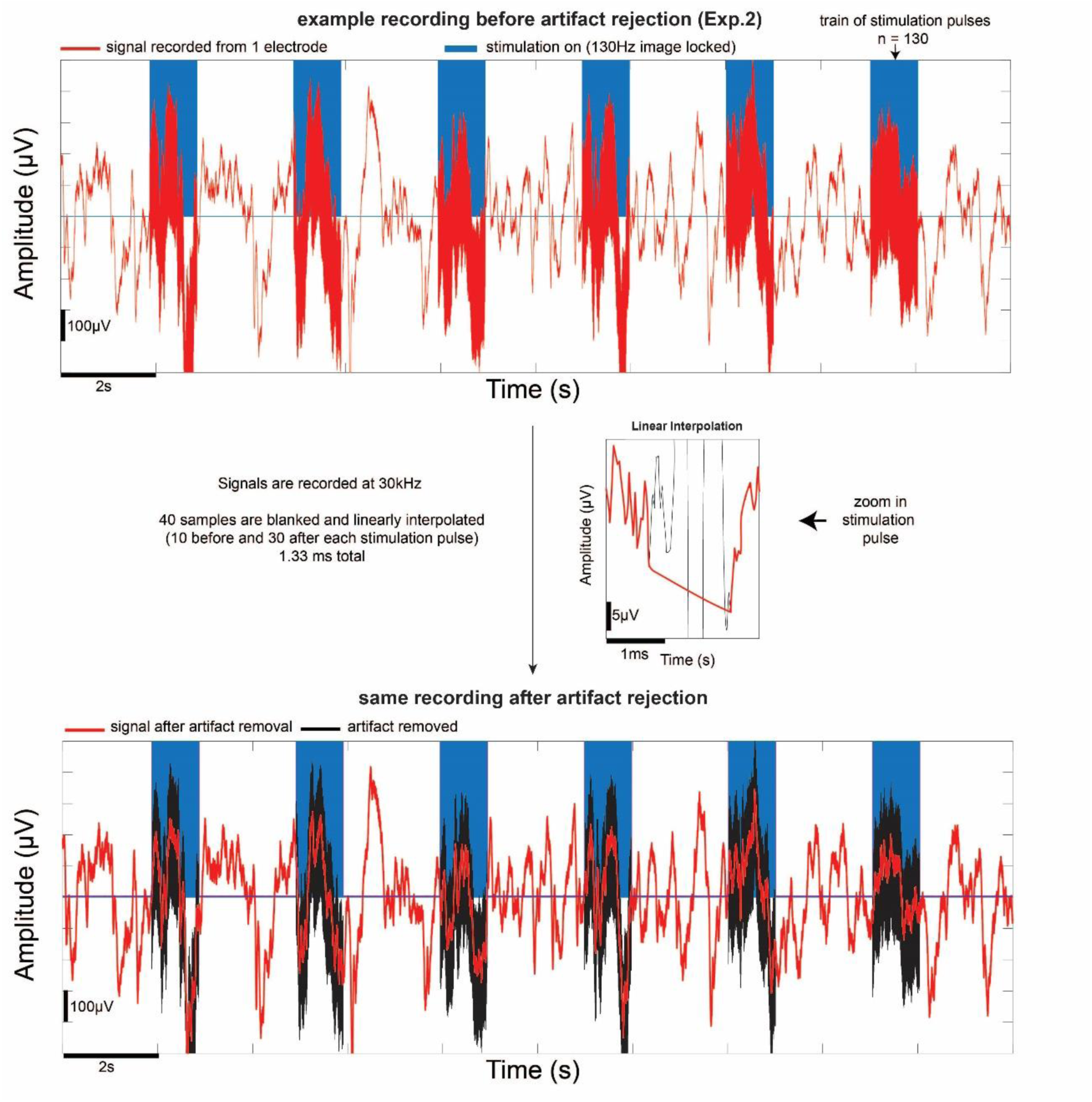
Blanking and interpolation of neural data during simulated experiments. Example recording from Exp.2 affected by stimulation artifacts (stimulation on depicted by blue squares) and the same recording after artifact rejection (original data before artifact removal shown in black for reference).

### Other locations with a significant modulation of time-frequency power values

In order to select the neuromodulation target, we performed an explorative time-frequency analysis of the signal from each recorded electrode during Exp.1 (baseline 1) and identified locations displaying increased spectral power to provocative images (versus neutral, p<0.05 using 5,000 permutations: see *Data-driven selection of stimulation location*). Three electrode locations within the rIFG displayed a significant difference, with significantly higher power in the 20-60 Hz range during provocative trials, 250-500 ms following go or no-go cue onset. For the experiments paired with neuromodulation, we selected one of the rIFG locations for stimulation (Figure 2 and Figure S2, top left panel) using a bipolar configuration with the adjacent rIFG location (Figure S2, middle top panel). Below, we report more details on the other locations displaying a difference in time-frequency power values depending on the image type. One location in the left dorsolateral prefrontal cortex (DLPFC) displayed higher power values in the 20-50 Hz range between 250-750 ms cue onset when exposed to provocative trials. One location in the left putamen also displayed significant differences around 250-750 ms following cue onset, in a lower frequency range (15-30 Hz). Finally, a location in the left hemispheric white matter, compatible with the location of the left arcuate fasciculus, generated significantly higher power values in trials with provocative images in the 10-40 Hz range, and these occurred 400-800 ms following cue onset. We also note that two locations (one in rIFG and the white matter location) displayed differences before the GNG cue onset, during the image presentation itself.

The same analysis was repeated for Exp.4 (Figure S3). The locations that displayed differences in Exp.1 no longer did for Exp.4, with no time-frequency power values surviving our statistical threshold in any of the locations. Thus, not only the rIFG difference was no longer present after neuromodulation, but this finding was true for all other locations as well (DLPFC, putamen and white matter).

**Figure S2.**
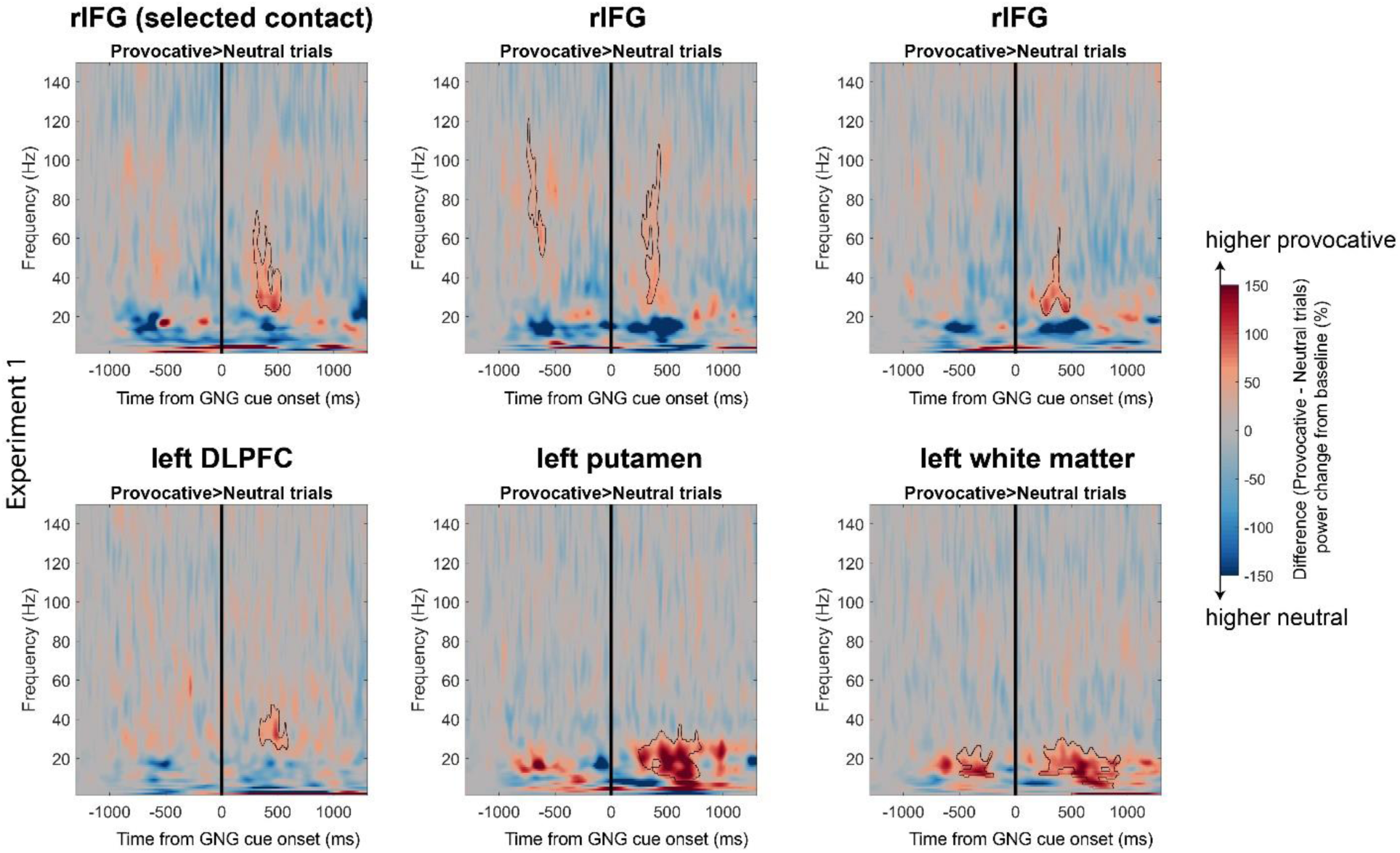
All contacts that displayed significantly greater power (% change from baseline) when exposed to provocative stimuli relative to neutral stimuli in Exp.1. The time-frequency maps represent the difference between the mean values for provocative versus neutral trials, and the presence of statistically significant increases for provocative trials are represented by black outlines (p<0.05, permutation testing with cluster-based correction for multiple comparisons).

**Figure S3.**
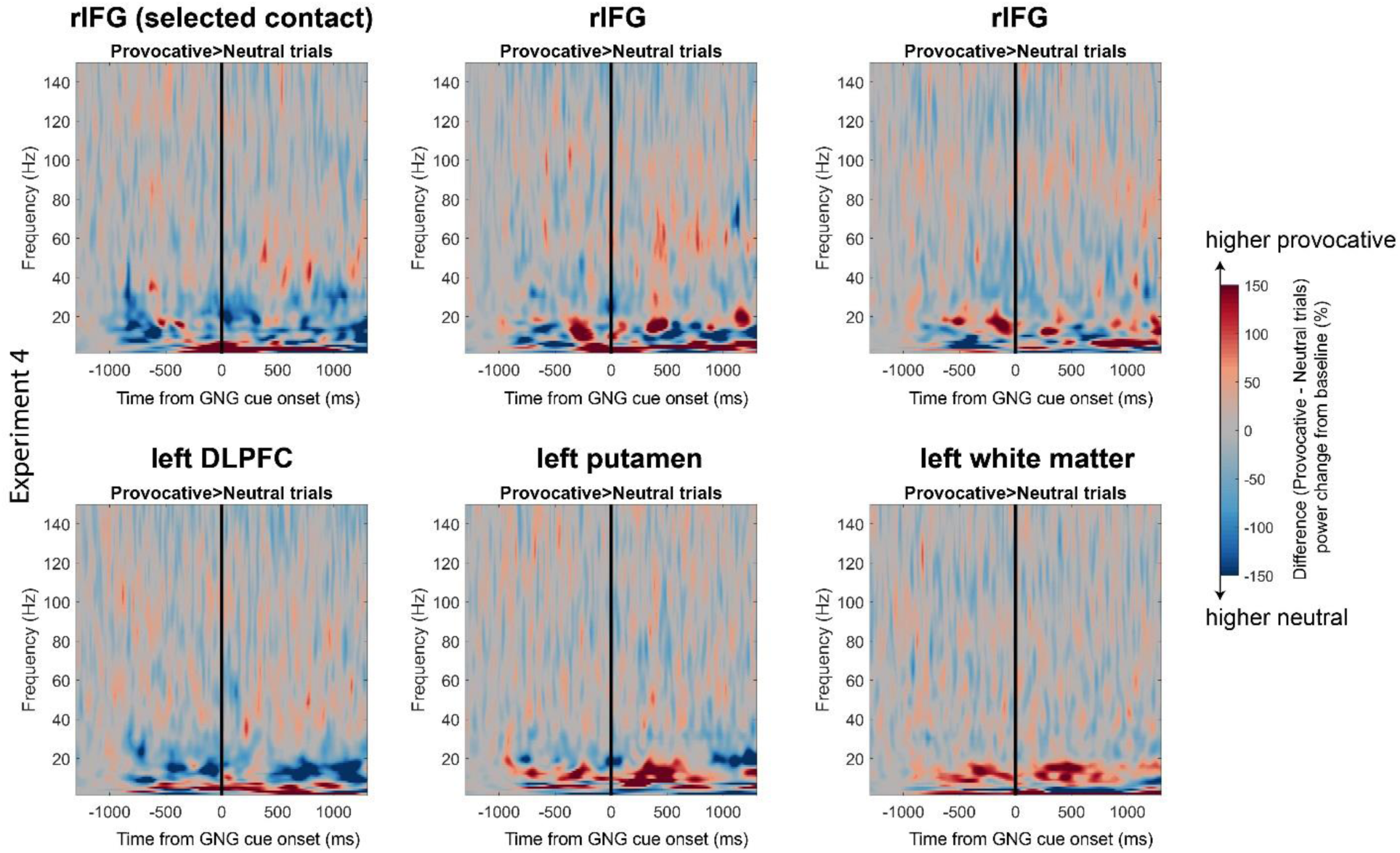
Same as S2 but for Exp.4. All contacts that displayed significantly greater power (% change from baseline) when exposed to provocative stimuli relative to neutral stimuli in Exp.1 were reassessed in the second baseline, after neuromodulation. No statistically significant increases for provocative trials were identified in any location (all p>0.05).

### P300 across all locations (not constrained by Exp.1)

In the main manuscript, we focused on the locations exhibiting a P300 in the first experiment and we evaluated changes in the P300 at these locations following neuromodulation. Thus, we constrained our analysis to the electrode locations present in Exp.1, and discarded additional locations displaying a P300 uniquely to subsequent experiments. Figure S4 displays all locations where an P300 was detected, even if not present in the first experiment. The same overall pattern is present when considering all the locations: the percentage of locations displaying modulations (normalized by the number of locations displaying a P300) is higher during low-performance experiments (Exp.1 and 2).

**Figure S4.**
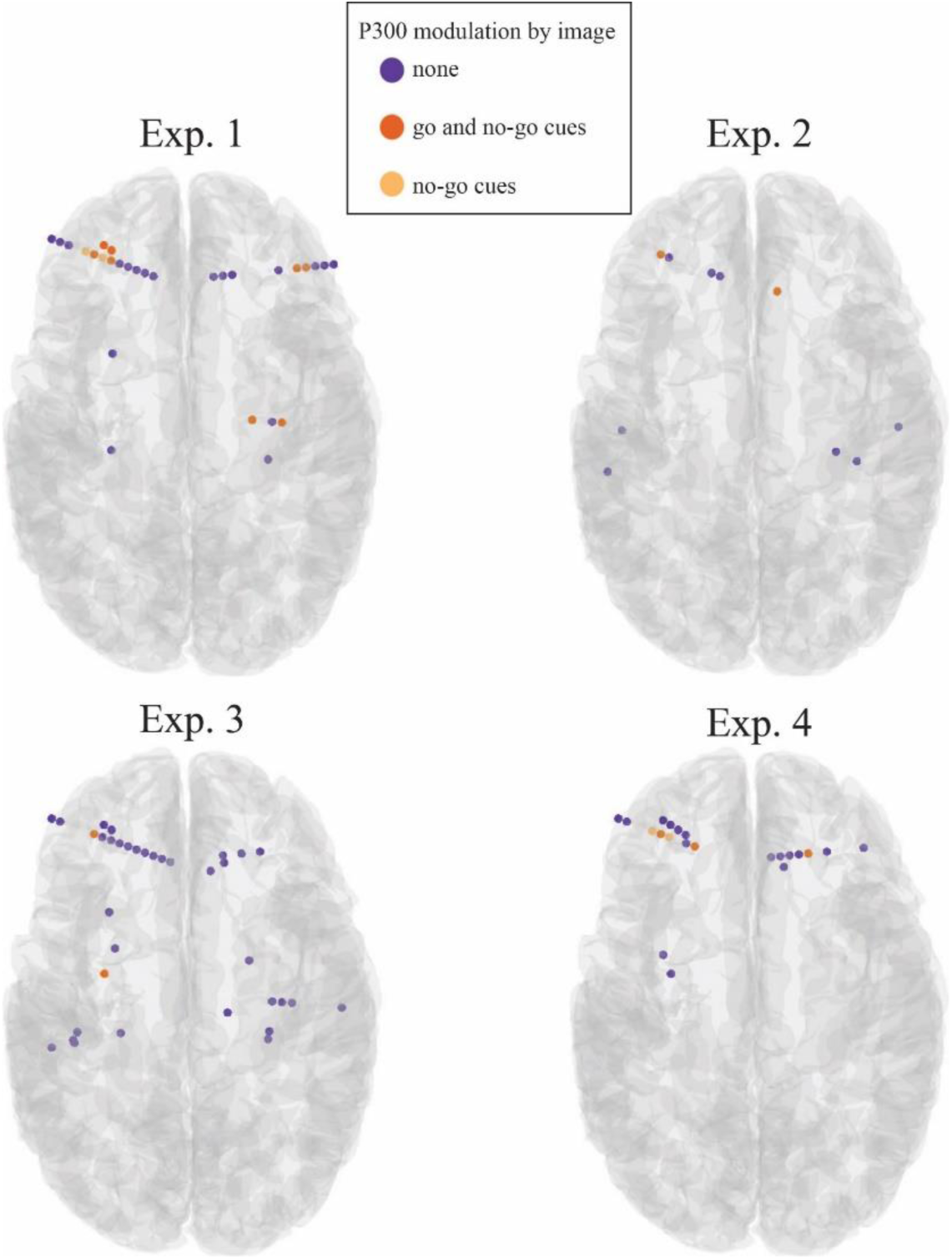
All locations displaying a P300 for each experiment.

